# Estimating the carcinogenesis timelines in early-onset versus late-onset cancers and changes across birth cohorts

**DOI:** 10.1101/2025.08.25.25334404

**Authors:** Navid Mohammad Mirzaei, Wan Yang

## Abstract

Understanding the timing of key mutational events in cancer development is critical for informing cancer prevention and detection strategies, particularly for early-onset cases that have increased in recent years. Yet intermediate mutational events are challenging to observe in humans. Here, we extend a tumor kinetic model we recently developed and long-term cancer registries data to estimate the expected timing of intermediate mutational events for breast, colorectal, and thyroid cancers. We formulate three distinct systems of ordinary differential equations, each describing cell evolution sequentially through three stages of carcinogenesis, up to the first occurrence of malignant transition. We further apply a convolution-based method to derive probability distributions and compute the expected age for each transition, based on parameters fit to incidence and tumor size data for each cancer. The models estimate the initial mutation occurs early in life for all three cancer types. For breast and colorectal cancers, estimated malignant transformation occurs more than a decade faster and hence earlier in early-onset than late-onset cases (in late 30s vs. late 40s to early 50s). In contrast, early- and late-onset thyroid cancers show similar early timelines (malignant transformation in late 20s), consistent with known early-life thyroid clonal activity. We also quantify early-onset carcinogenesis timelines for three key birth cohorts (born during 1950-1954, 1965-1969, and 1980-1984) and identify a shift toward earlier malignant transformation in more recent cohorts, largely due to faster progressions of later stage transitions, for all three cancer types. These findings can inform early-onset cancer etiologic studies and intervention strategies.

**Significance Statement:** Carcinogenic mutational events are crucial for cancer etiology but challenging to observe. Here, we circumvent the research challenges to trace key mutations stepwise through the emergence of malignancy by combining long-term cancer registry data and tumor kinetic modeling for three key cancer types (breast, colorectal, and thyroid). We find early-life initial mutations but diverging timelines in cancer progression between early- and late-onset cases and temporal changes across birth cohorts. We estimate faster progression and hence earlier onset in more recent birth cohorts, consistent with the reported increases in early-onset cancer incidence. Our study provides the first direct, cohort-specific estimates on when key mutational transitions occur over the life course, which can inform early-onset cancer etiologic studies and intervention strategies.

## INTRODUCTION

Most cancers develop through a multistep process, where cells accumulate genetic alterations before the emergence of malignancy. Understanding when these intermediate transitions occur is critical for guiding early prevention and detection strategies. This is particularly relevant and urgent for early-onset cancers (i.e., diagnosed under age 50). Recently, incidence of multiple cancer types (e.g., breast and colorectal cancer) has risen among young adults and in more recent birth cohorts, and the causes remain largely unclear [1]. A better understanding of changes in intermediate carcinogenic timelines in early-versus late-onset cancers and changes across birth cohorts can help reveal the reasons behind the early-onset cancer increases and inform public health intervention strategies.

Yet, studies investigating the timing of intermediate mutagenic events remain limited. The seminal multistage carcinogenesis model was built on the multistep carcinogenesis theory [2]. Later multistage clonal expansion (MSCE) models further incorporated mutation and proliferation dynamics to describe progression from normal cells to malignancy [3]. These kinetic models have helped explain how age-specific incidence patterns emerge across cancer types. Nonetheless, such models typically focus on the malignant transition (i.e., emergence of the first cancer cell) or the sojourn time (i.e. interval) between malignant transformation and clinical detection, and do not estimate the timing of intermediate carcinogenic transitions [4-7]. More recently, Lahouel et al. developed a detailed multistage tumor growth model at the individual level, which incorporates typical carcinogenic dynamics (e.g., birth, death, mutation processes) plus fitness advantages from specific driver mutations [8]; they estimated the timelines of key driver mutations and suggested the first driver event generally occurred at early ages. Gerstung et al. used whole-genome sequencing of over 2000 tumors to model the mutational and evolutionary history of 38 cancer types [9]; similarly, they conclude that tumorigenesis often begins much earlier than traditionally assumed. However, these estimates are either partially calibrated to data (e.g., 4 out of 19 model parameters in [8] were estimated using incidence data and only for colorectal cancer) or remain coarse (e.g., some estimates spanned decades in [9]). Further, none of these studies focused on early-onset cancers.

In a recent work, we developed an MSCE-T model (T for tumor), extending the classic MSCE framework, to incorporate tumor growth from malignant transformation to detection [4]. Assuming there are three key mutational transitions per Tomasetti et al. [10], this model included sequential transitions from normal stem cells to First Stage Mutated Cells (FSMCs), Second Stage Mutated Cells (SSMCs), and ultimately malignant cells up to the time of tumor detection (see Figure 1). In [4], we prove the MSCE-T model is fully identifiable, and showed that it better distinguishes between true increases in cancer risk and earlier detection (a contending hypothesis) for three key early-onset cancers with increased incidence – breast cancer (BrC), colorectal cancer (CRC), and thyroid cancer (ThC).

**Figure 1.**
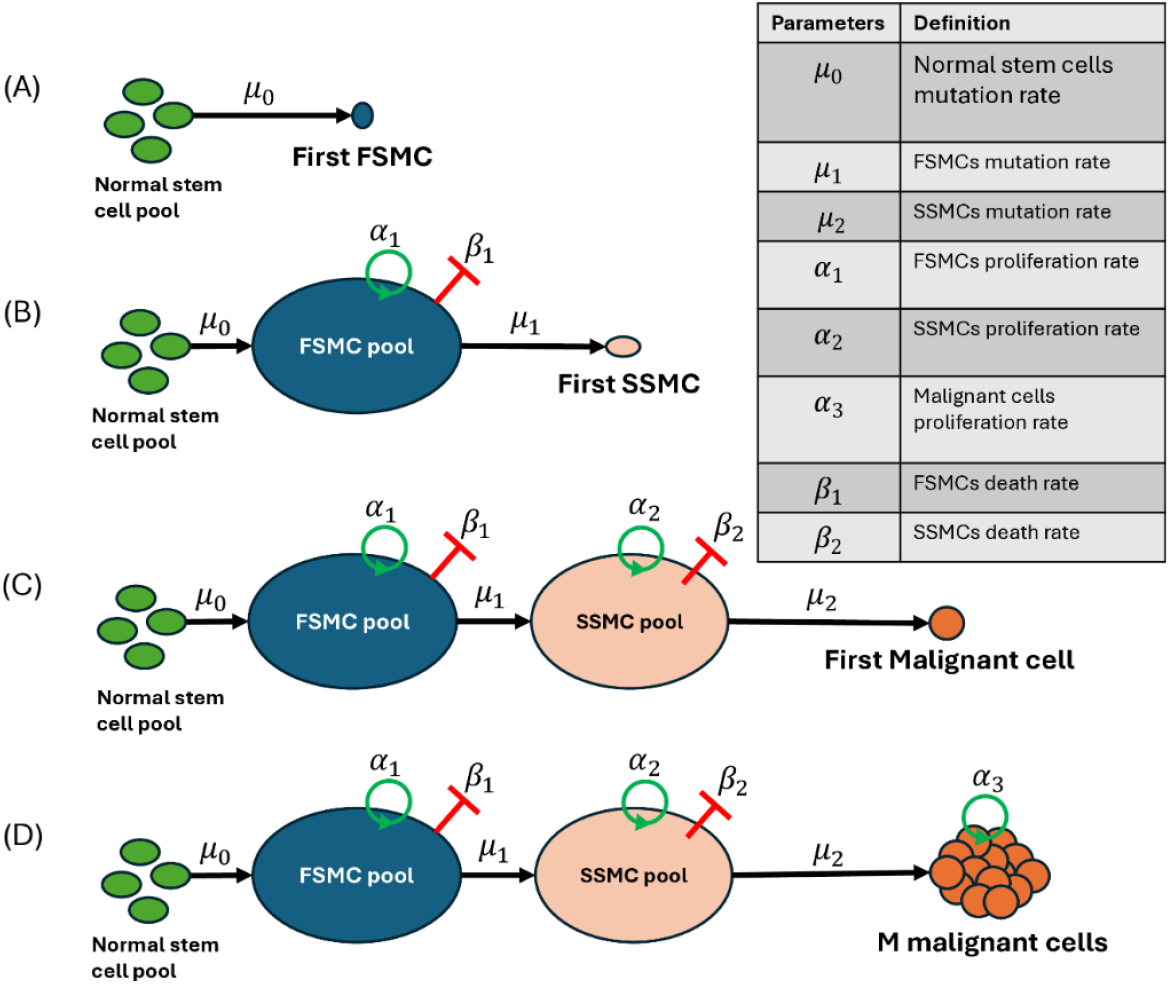
The schematics of 4 model classes. (A) Normal stem cells acquire a first mutation at rate *μ*_0_, generating the first First-Stage Mutated Cell (FSMC); the model outputs the incidence of first FSMC appearance. (B) Extending (A), this model includes proliferation (α_1_), death (*β*_1_), and mutation (*μ*_1_) of FSMCs to Second-Stage Mutated Cells (SSMCs), outputting the incidence of first SSMC occurrence. (C) Building on (B), SSMCs proliferate (*α*_2_), die (*β*_2_), or mutate into malignant cells at rate *μ*_2_; the model outputs the incidence of the first malignant cell. (D) The MSCE-T model from our work in [4], which further extends (C) by including malignant cell proliferation at rate *α*_3_. This model outputs cancer incidence based on the malignant cell population reaching the reported tumor size at diagnosis.

Here, we leverage the modular structure of the MSCE-T model [4] to isolate each mutational transition timing, using convolution-based analysis of probability distribution functions (PDFs). We derive three ordinary differential equation systems, each capturing the emergence of FSMCs, SSMCs, or malignant cells. We estimate MSCE-T model parameters with cohort- and age-specific cancer incidence data, and further use those parameters and the derived PDFs to estimate the timelines for all three transitions in breast, colorectal, and thyroid cancers, for both early-onset (under age 50) and late-onset (through age 70) cases. To our knowledge, this is the first study to estimate intermediate mutation timelines using an identifiable cancer kinetic model (here, the MSCE-T model) based on cohort- and age-specific cancer incidence data.

## MATERIALS AND METHODS

### Model framework

Our models are based on the survival probability S(t) = Pr(t < T), where T is a random variable denoting the time of an adverse event. Let F(t) be the cumulative distribution function for Pr(t ≥ T), then we have:

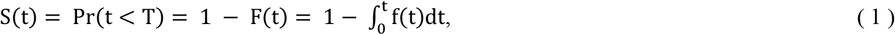

where f(t) is the PDF associated with F(t). Then, the hazard function associated with *S*(*t*) is defined as:

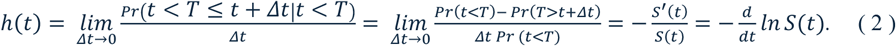

To track the timing of the three intermediate mutational events illustrated in Figure 1 (A)-(C), we utilize three survival probability sets. They are defined as the probability of: (A) having no FSMCs; (B) having no SSMCs; and (C) having no malignant cells. Denoting the number of normal stem cells, FSMCs, SSMCs and Malignant cells at time t as I_0_(t), I_1_(t), I_2_(t) and M(t), respectively, and letting I_0_(t) = N_0_ for all time given the pool size of normal stem cells tends to remain constant [11] (with *N*_0_values for each cancer taken from [12, 13]), the fundamental survival probabilities we use to derive the systems are given below:

For class (A):

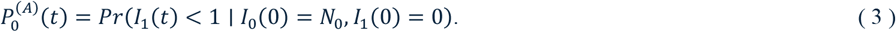

For class (B):

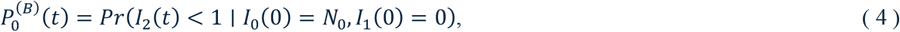

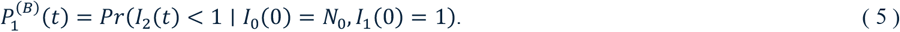

For class (C):

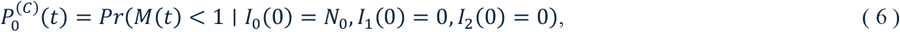

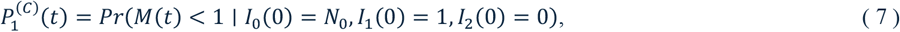

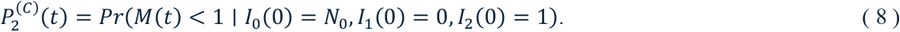

The superscripts correspond to three classes listed above and in Figure 1 (A)-(C). Based on survival probabilities Eqs.3-8 and the mutational transitions illustrated in Figure 1 (see parameter notations in the table inset), we get the following equation systems (see the derivation for all three model classes in the supplementary materials).

For class (A):

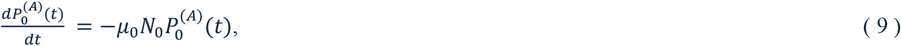

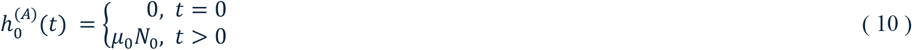

Here,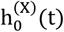 is the hazard calculated via Eq. 2 for the survival probability 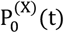, where (X) is (A), (B), or (C) depending on the model’s class. However, for class (A), Eq.2 yields a constant hazard, so we set 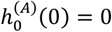 to prevent mutational events at time zero. The initial condition for Eq.9 is 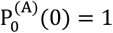.

For class (B):

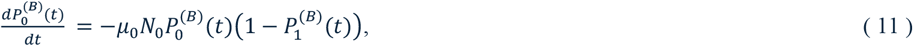

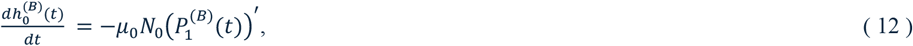

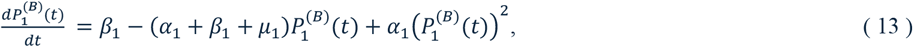

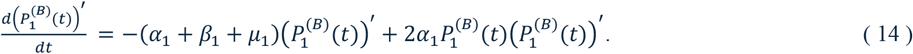

Here, ()′ means derivative with respect to t and 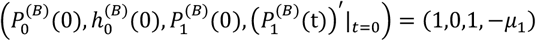.

Finally, for class (C):

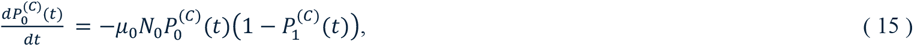

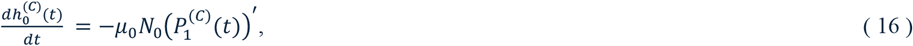

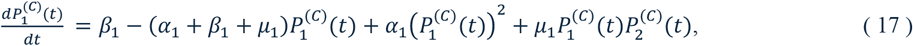

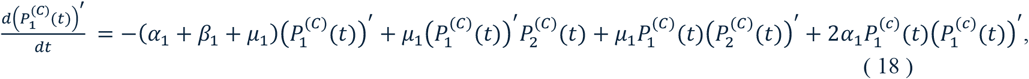

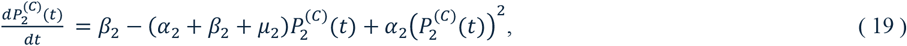

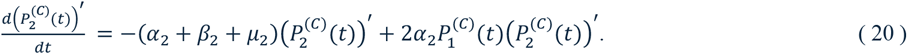

Figure 1 (D) illustrates the full MSCE-T model developed in our previous study [4]; see [4] for a detailed derivation, which further entails the use of a moment-generating probability density equation to obtain a closed-form expression for *P*_*n*_ (*t*), in addition to similar steps used to derive models (A)-(C).

### Calculating the timing of different mutations

The appearance of the first FSMC through normal stem cells mutation is independent of the SSMCs and malignant cells, yet FSMCs are precursors of and thus essential to the other two groups. Therefore, we start with the timing of the first FSMC emergence. From model (A), we can calculate the survival and hazard functions and using Eq.1 and Eq.2 we can find the associated PDF:

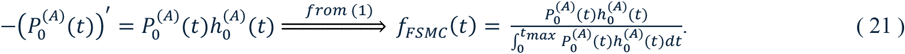

Note, here the survival function S(t) in Eq.1 and Eq.2 is replaced by the model specific survival 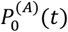. Dividing by the integral in Eq.21 ensures the PDF is proper (i.e., the area under its curve from 0 to the maximum simulation time *t*_max_ is 1). Function *f_FSMC_*(*t*) quantifies the probability of FSMC happening at exactly age *t*, given it has yet to happen. The expected time for this occurrence is thus:

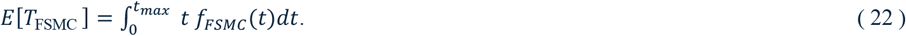

Estimating the timing of SSMC and malignant cell appearance is more complex, as their emergence depends on the prior occurrence of mutated cells from the preceding stage. However, our model’s hierarchical structure enables derivation of the marginal probability distributions of downstream transitions using those of upstream events. For _1_ example, for SSMCs, we use the survival probability 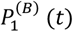 from model (B), to determine the time distribution of the first SSMC appearance, conditional on prior FSMC emergence:

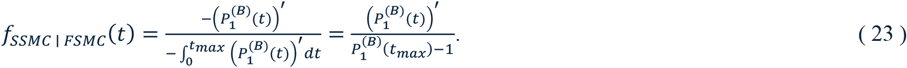

Then using PDF convolution with Eq.21, we get

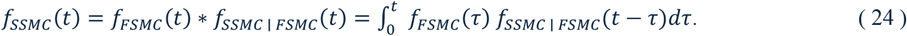

and its expectation:

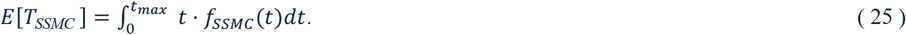

Similarly, from model (C) we get:

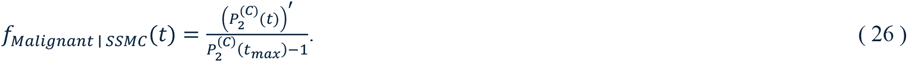

Using convolution with Eq.25 (note, it already considers the occurrence of FSMCs), we can get the PDF for malignant cells:

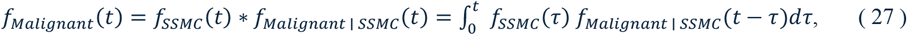

and its expectation:

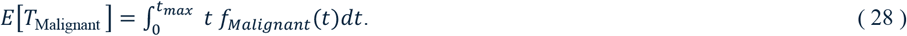

### The workflow and analyses

Figure 2 outlines the workflow of this study. Model (D) is first fit to cancer incidence data from the Surveillance, Epidemiology, and End Results (SEER) program [14] for three cancer types (i.e., BrC, CRC and ThC), as done in [4]; see supplementary materials for a summary. Importantly, we note the MSCE-T model (i.e., Model (D)) explicitly models tumor growth from the first malignant cell to tumor detection based on tumor-size-at-diagnosis data; in doing so, the parameter estimates here account for the time interval (i.e., sojourn time) from first malignancy to tumor detection as well as changes in tumor detection over time (e.g., earlier detection with smaller tumor size in more recent years). In addition, for BrC, we incorporate an initial delay to account for the inactive state of normal mammary stem cells before puberty [15], based on menarche age (see supplementary materials). The estimated parameters are then fed to models (A), (B) and (C). Using the outputs from these models we can directly calculate f_FSMC_(t), f_SSMC | FSMC_(t), and f_Malignant | SSMC_(t), per Eqs.21, 23, and 26, respectively. Then using PDF convolution, we evaluate f_SSMC_(t) and f_Malignant_ (t) per Eqs.24 and 27, respectively. Finally, the expected first occurrence time for FSMCs, SSMCs, and Malignant cells can be calculated per Eqs.22, 25, and 28, respectively.

**Figure 2.**
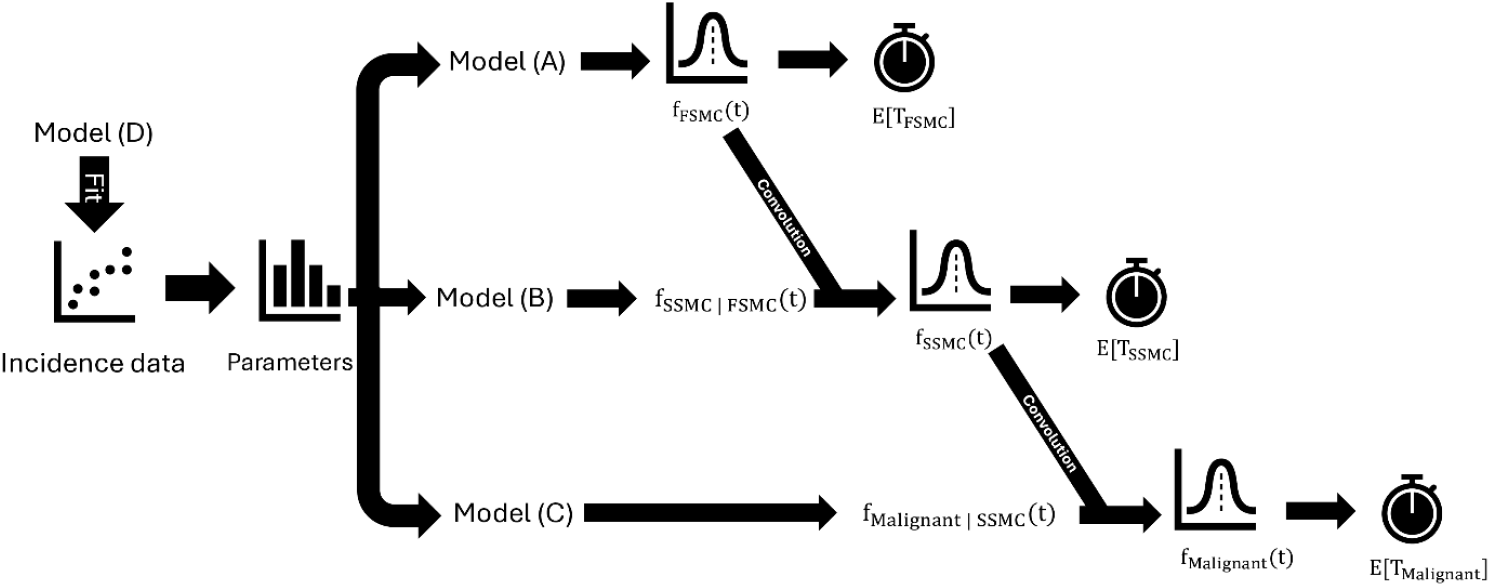
Study workflow. Parameter estimates are first obtained by fitting model (D) to cohort- and age-specific cancer incidence data. The estimated parameters are then fed to Models (A), (B) and (C). The PDF associated with the emergence of FSMCs can be directly evaluated from model (A)’s outputs, while for the emergence of SSMCs and Malignant cells a convolution is carried out per Eq.24 and Eq.27. From the PDFs, we can evaluate the expected time of each cell type’s occurrence.

In our first analysis, we estimated the timelines for the 1945–1949 birth cohort for BrC, CRC and ThC, for cancers diagnosed i) under age 50 and ii) through age 70, to compare the early-onset versus late-onset cases. We chose the 1945-1949 cohort, as cancer-screening coverage was low in this early cohort––under 30% for BrC and 34% for CRC [16, 17], allowing us to perform parameter estimation through age 70 without explicitly modeling the impact of screening on cancer incidence.

In the second analysis, we focus on early-onset cancers, for three representative cohorts born in 1950-1954, 1965-1969, and 1980-1984 separately. In the U.S., BrC and CRC have established screening guidelines; specifically, the U.S. Preventive Service Task Force (USPSTF) recommended screening starting at age 40 for BrC and age 50 for CRC prior to 2021 (i.e., applied to our study period) [18, 19]. Thus, to estimate early-onset carcinogenesis timelines and minimize screening-related biases, we fit the MSCE-T model to cohort-specific data for cases diagnosed under age 40 for BrC and under age 50 for CRC. For comparison, we similarly limited the model fitting to incidence under age 50 for ThC (no screening in the U.S.).

## RESULTS

### Cancer Incidence Trends and Model Fits

Figure 3A shows incidence data for individuals born in 1945-1949 and model fits using the MSCE-T model. For early-onset (under age 50; Figure 3A top row) and late-onset (through age 70; Figure 3A bottom row), the MSCE-T model accurately reproduces the incidence trends, demonstrating its capability to capture the age-specific cancer incidence. As noted in the Introduction, early-onset cancer incidence has increased. This is evident for all three cancers examined here. As shown in Figure 3B, incidence rates of BrC among those under age 40 (note: screening for BrC starts at age 40, hence we cap the age limit at 40), and rates of CRC and ThC among those under age 50 have all increased in more recent birth cohorts (see darker colors for more recent cohorts in Figure 3B). The MSCE-T model also closely reproduces these cohort-specific trends (Figure 3B).

**Figure 3.**
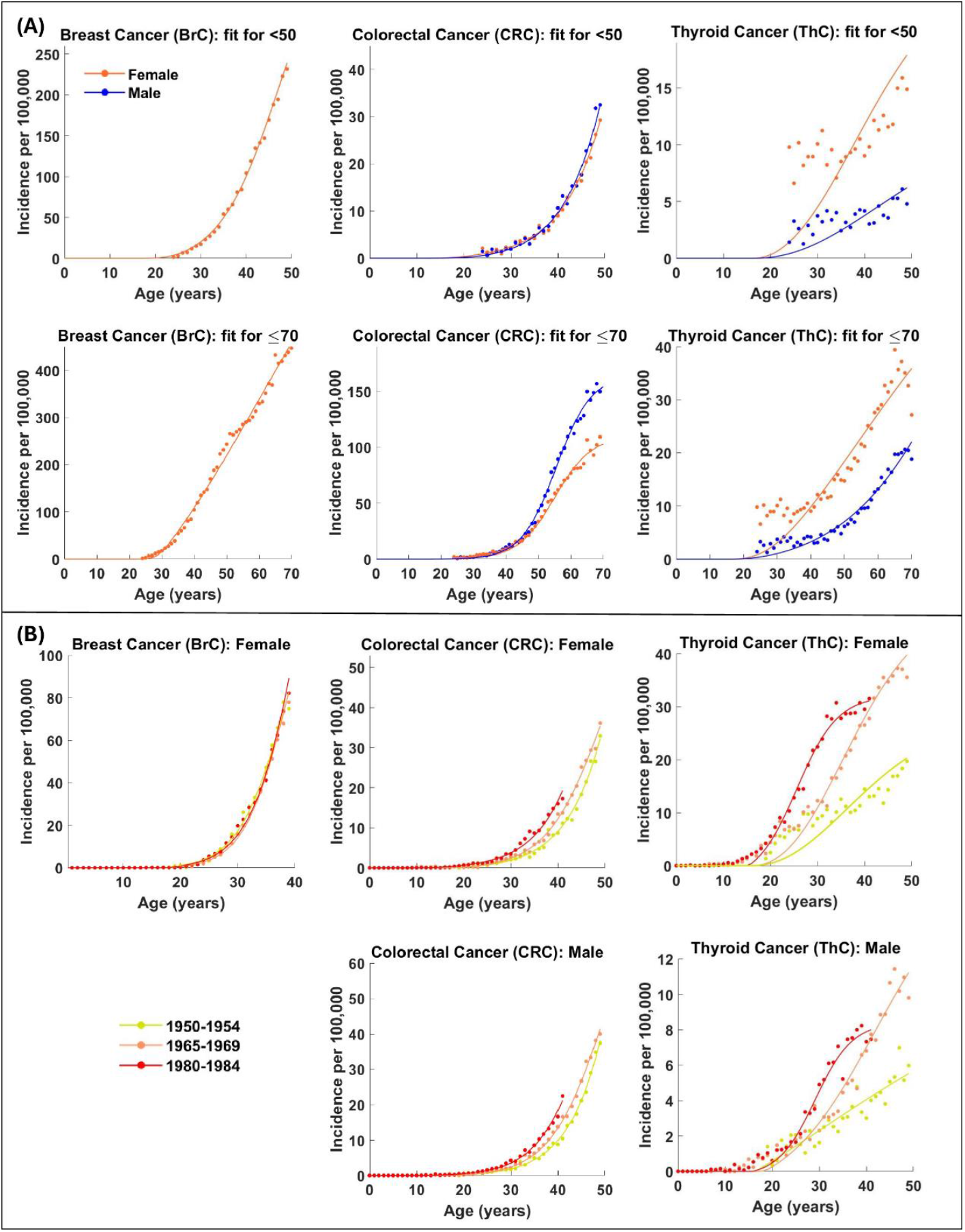
MSCE-T model fits for three cancer types. (A) MSCE-T model fits for the 1945-1949 birth cohort, for early-onset cases diagnosed before age 50 (top row) and late-onset cases through age 70 (bottom row). (B) MSCE-T model fits for incidence of three cohorts born in 1950-1954, 1965-1969, and 1980-1984. The model fits (lines) are generated using SEER data (shown by the dots) for cases diagnosed from 1973-2021; note calendar year equals birth year (color coded; see legend) plus age (see x-axis).

### Early-Onset vs Late-Onset Mutational Timelines in the 1945–1949 Cohort

Table 1 compares early- and late-onset mutational timelines estimated for the 1945–1949 cohort. These include the mean ages at three key transitions: i) Normal → FSMC, ii) FSMC → SSMC, and iii) SSMC → Malignant (Figure 1). For BrC and CRC, the primary difference appears to be in the final transition to malignancy, with much earlier mean age of malignant transformation estimated for early-onset (under age 50) than late-onset (through age 70) cases in this cohort. For female BrC, estimated mean age of transition from SSMC to first malignancy was 39.5 years among early-onset cases, compared with 52.9 years among late-onset cases. Similarly, for CRC, this transition generally occurred 10+ years earlier among early-onset cases for both sexes (early-vs late-onset: 38.3 vs. 50.9 years in females, and 37.8 vs 47.8 years in males). The two preceding transitions (Normal → FSMC and FSMC → SSMC) also occurred slightly earlier in early-onset cases, as shown in Table 1.

**Table 1.** Comparison of early-onset versus late-onset mutational timelines for breast, colorectal, and thyroid cancers, estimated for the 1945–1949 cohort. The early-onset timelines are estimated using incidence data limited to ages <50, whereas late-onset timelines are estimated using incidence data through and including age 70.

| Cancer Type | Sex | Type | Mean Age:<br>Normal→FSMC (in<br>years) | Mean Age:<br>FSMC→SSMC<br>(in years) | Mean Age:<br>SSMC→Malignant (in<br>years) |
| --- | --- | --- | --- | --- | --- |
| Breast cancer (BrC) | Female | *Early-onset (<40) | 13.0 (CI: 12.9-13.1) | 35.0 (CI: 34.9-35.1) | 37.3 (CI: 37.2-37.4) |
|  |  | Early-onset (<50) | 14.2 (CI: 14.0-14.3) | 36.6 (CI: 36.5-36.7) | 39.5 (CI: 39.4-39.6) |
|  |  | Late-onset (≤70) | 14.0 (CI: 13.9-14.2) | 39.0 (CI: 38.8-39.2) | 52.9 (CI: 52.7-53.1) |
| Colorectal cancer (CRC) | Female | Early-onset (<50) | 4.6 (CI: 4.6-4.7) | 36.5 (CI: 36.4-36.5) | 38.3 (CI: 38.2-38.4) |
|  |  | Late-onset (≤70) | 4.6 (CI: 4.5-4.7) | 36.7 (CI: 36.7-36.8) | 50.9 (CI: 50.9-51.0) |
| CRC | Male | Early-onset (<50) | 3.9 (CI: 3.9-4.1) | 32.1 (CI: 32.0-32.2) | 37.8 (CI: 37.7-37.9) |
|  |  | Late-onset (≤70) | 4.3 (CI: 4.2-4.4) | 33.0 (CI: 33.0-33.1) | 47.8 (CI: 47.7-47.9) |
| Thyroid cancer (ThC) | Female | Early-onset (<50) | 4.5 (CI: 4.4-4.6) | 14.0 (CI: 13.9-14.1) | 26.9 (CI: 26.8-27.0) |
|  |  | Late-onset (≤70) | 4.4 (CI: 4.3-4.4) | 16.7 (CI: 16.5-16.8) | 27.4 (CI: 27.3-27.6) |
| ThC | Male | 1945-1949 (<50) | 2.0 (CI: 1.9-2.1) | 17.7 (CI: 17.6-17.9) | 26.8 (CI: 26.7-26.9) |
|  |  | 1945-1949 (≤70) | 2.1 (CI: 2.0-2.2) | 20.8 (CI: 20.7-20.9) | 27.0 (CI: 26.9-27.1) |

\*This row is added for comparison with Table 2 estimates for BrC <40.

In contrast, for ThC, we estimate similar early timelines regardless of the age cut-off used (i.e., diagnosed under age 50 vs. through age 70; Table 1). For both sexes, estimated initial mutation occurred in early childhood (∼2-5 years of age) and the estimated mean age at malignant transition was ∼27 years for both sexes, using either age cut-off (Table 1). The main difference – by ∼3 years – appears to be in the second transition (FSMC → SSMC) – we estimate this transition occurred during teenage years (∼14-17) for females and early adulthood (∼18-21) for males, for the 1945-1949 cohort. These early lifetime estimates are consistent with reports from the literature; for example, see Figure 2 of Takano 2017 [20], where detailed autopsy data showed the prevalence of ThC increases steeply from age 15 to 34 and then stays roughly constant for the remaining lifetime.

### Trends in Early-Onset Carcinogenesis Across Birth Cohorts

Table 2 compares the estimated early-onset mutational timelines for BrC, CRC, and ThC among three representative birth cohorts over a timespan of 30 years: those born during 1950-1954, 1965-1969, and 1980-1984. For early-onset BrC, estimated mean age at which the first mutation occurs (i.e., Normal → FSMC) consistently aligned with the age of menarche (∼13 years; see Table S1) for all three cohorts. That is, the first steps in BrC appear to occur almost immediately following the onset of breast development. However, the later, more advanced transitions (i.e., FSMC → SSMC, and SSMC → Malignant) appear to occur earlier in more recent cohorts. Estimated mean age at the 2^nd^ transition (FSMC → SSMC) reduced by ∼2.5 years in cohorts born 30 years apart (mean age: 34.8 years in the 1950-1954 cohort vs. 32.4 years in the 1980-1984 cohort) and estimated mean age of malignancy reduced by ∼1 year (37.2 years vs. 36.0 years in those two cohorts).

**Table 2.**
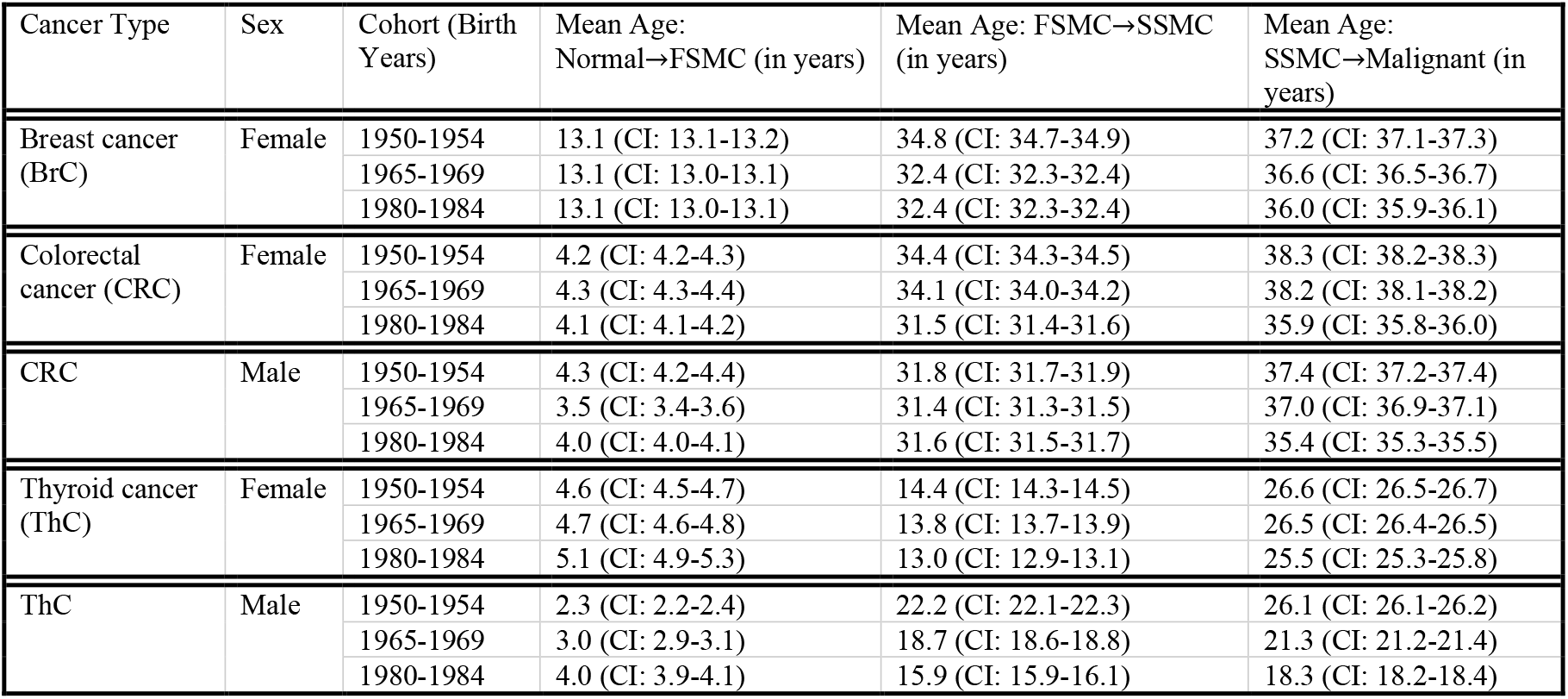
Comparison of mutational timelines across birth cohorts, for early-onset breast, colorectal, and thyroid cancers. For this analysis, we generate the estimates for breast cancer using incidence data for ages <40, as breast cancer screening starts at age 40. For colorectal and thyroid cancers, we generate the estimates using incidence data for ages <50.

For CRC, our models consistently estimate every initial mutation, at ∼4 years of age, for both early-onset and late-onset cases, and all sexes and cohorts (Tables 1 and 2). However, the models estimate faster and thus earlier late-stage transitions, particularly the malignant transformation in more recent cohorts. In addition, such accelerations were more substantial in females born during 1980-1984, the latest birth cohorts in this study (Table 2); estimated mean age reduced by ∼3 years (from 34.4 to 31.5 years of age; birth cohorts 1980-1984 vs 1950-1954) and by ∼2 years (from 38.3 to 35.9 years of age in those two cohorts) for the last two transitions, respectively, compared to females born 30 years earlier. Estimated mean age of malignant transition in males born during 1980-1984 was also ∼2 years younger than those born 30 years earlier (mean age: 35.4 vs. 37.4 years), but the 2^nd^ transition remained stable across cohorts in males (∼31-32 years; Table 2).

For early-onset ThC, our models estimate pronounced cohort-dependent changes, especially for males. As noted above, ThC tends to develop early in life and remain indolent [20, 21]. Despite this early carcinogenesis timeline, our models estimate significant shifts toward earlier late-stage transitions for males in more recent cohorts, by ∼6 years for the FSMC → SSMC transition and by ∼8 years for the SSMC → Malignant transition, comparing the 1950-1954 and 1980-1984 cohorts (Table 2); estimated shifts were less pronounced among females across the three cohorts. However, caution is advised when interpreting the results for ThC since the model is unable to mimic the initial incidence data with the same precision as the other two cancers (see Figure 3).

Overall, our models consistently estimate early initiations, with the first mutational event occurring during early childhood for both CRC and ThC and immediately following breast development for BrC; these findings align with previous reports [8, 9]. The models also indicate a shift toward faster and earlier malignant transformation in more recent birth cohorts, for all three types of early-onset cancer (i.e., BrC, CRC and ThC).

## DISCUSSION

Early-onset cancer incidence has increased rapidly in recent years, prompting discussion on whether this trend reflects earlier detection or changes in risk exposures. To investigate this, here we focus on two key unknowns: i) how early-onset cases differ from late-onset cases in the timing of key mutational events; and ii) whether there is a generational shift in the carcinogenesis timeline, including the age at which each mutation occurs and the duration between tumor initiation and malignant transformation. As observation is not feasible, we circumvent the research challenges by combining long-term cancer registries data and tumor kinetic modeling. Tracing the mutations stepwise, our study provides the first direct, cohort-specific estimates on when key mutational transitions occur over the life course, for three key cancers (BrC, CRC, and ThC). We estimate early initial mutations for all three cancer types but diverging timelines in cancer progression between early- and late-onset cases and temporal changes across birth cohorts. These findings offer new biological insights that can inform more in-depth etiologic studies and cancer intervention strategies.

Our analysis of early-versus late-onset cases in the 1945-1949 cohort suggests that early-onset BrC and CRC cases progress more rapidly than late-onset cases, mostly due to much faster progression in the malignant transition. With relatively similar estimates for the mean age at initiation and mean age at the 2nd transition (FSMC → SSMC), early-onset cases progress to malignancy 10+ years earlier than late-onset cases. For example, for female CRC, from the appearance of the first SSMCs to malignant transition took ∼2 years for early-onset cases vs. ∼14 years for late-onset cases, leading to a much earlier mean age of malignancy (38.3 in early-onset cases vs. 50.9 in late-onset cases). A similar 10-year gap in the last transition time was estimated for male CRC and female BrC. These substantial differences suggest that the later stages of progression and a much-compressed timeline for the malignant transition step—rather than initiation—are primarily responsible for the earlier emergence of malignancy in early-onset cancers. This finding aligns with growing evidence that early-onset CRC and BrC have distinct mutational profiles compared to their late-onset counterparts and exhibit more aggressive behavior [22, 23].

Examining early-onset cases in three more recent cohorts of 1950-1954, 1965-1969 and 1980-1984, we find further accelerations in later stage mutational transitions for both BrC and CRC. For example, for early-onset female CRC, our model estimates the 2^nd^ transition took ∼3 years less, accelerating the mean age of occurrence to 31.5 years in the 1980-1984 cohort, from 34.4 years of age in the 1950-1954 cohort (Table 2). In contrast, the estimated mean initiation age remained consistent over the 30 years. These estimates, consistent with our 1^st^ analysis comparing early- and late-onset cases, suggest accelerations in later stage mutational transitions (as opposed to earlier initiations) may explain earlier onset in recent cohorts. It is challenging to pinpoint a cancer risk factor for early-onset cancers, due to the potential long latency [1] and limited early-life exposure data. Our estimates here can help design detailed studies to examine potential risk factors (e.g., obesity and alcohol consumption) that may drive the recent early-onset incidence increase, based on both the changes in exposure prevalence across cohorts and the windows of exposure (e.g., teenage years vs. mid-life). For instance, future work could incorporate exposure data to test which exposures align with the timeline shifts estimated here.

We also find earlier malignant transformation in more recent cohorts, for all three cancer types (Table 2). Such shifts in carcinogenesis timelines have direct implications for screening policy. For early-onset CRC, estimated mean age of malignant transformation declined from 38.3 (for females) and 37.4 years (for males) in the 1950-1954 cohort to 35.9 years and 35.4 years in the 1980-1984 cohort, respectively. For early-onset BrC, estimated mean age of malignant transformation also decreased by ∼1 year, to 36 years of age in the 1980-1984 cohort. These findings point to benefits of reevaluating age-based risk assessment and screening thresholds, given the rising early-onset incidence and accelerated carcinogenesis timeline. Similar concerns have recently led to updates on CRC screening guidelines recommending initiation at age 45 (vs. age 50 before 2021) [24], as well as European recommendations for high-risk individuals to begin annual breast MRI screening as early as age 25, with the addition of mammography starting between ages 35 and 40 [25].

Thyroid cancer is traditionally considered indolent [20, 21] and our results (for the 1945-1949 cohort) show similar timing of malignancy transition regardless of the age cut-off used for estimation. However, for later cohorts, our models estimate substantial cohort-dependent shifts, particularly among males. In men, estimated mean age at malignant transition dropped from 26.1 years in the 1950–1954 cohort to just 18.3 years in the 1980–1984 cohort—a ∼8-year drop. Importantly, this occurs despite the consistent estimated early initiation ages (∼2–4 years), suggesting accelerated late-stage progression in more recent cohorts. Some studies attribute rising ThC to overdiagnosis [26, 27], while others noted a true increase (e.g., based on stage-specific incidence data in Lim et al. 2017 [28]). Here, we account for changes in detection by incorporating reported tumor-size-at-diagnosis data [4], and our results support a real biological acceleration. The reasons behind such changes remain unclear, although several hypotheses have been proposed (see, e.g., Vigneri et al. 2015 [29]). In addition, the clinical implications of these results are nuanced. On the one hand, since most ThC are indolent and benign, the need for early detection and active intervention is open for debate. On the other hand, the rise of thyroid cancer incidence may indicate rising exposure levels (e.g., endocrine disruptors) that might also increase the risk of other cancer types. Thus, more in-depth analyses on lethal ThC subtypes (e.g., anaplastic ThC) as well as potential link between thyroid disfunction and other cancers are warranted.

This work utilizes the MSCE-T modeling framework and consequently shares its limitations. First, the three-stage carcinogenesis model applied uniformly across all three cancer types follows the structure proposed by Tomasetti et al. [10], which, while supported by prior evidence, may not capture full tumor evolution complexity in each context. Nonetheless, the estimated changes by birth cohort should hold regardless, as the same model structure is used for all cohorts. Second, we assume all model parameters remain constant over time, a simplification that does not account for potential biological variability or time-varying risk exposures. Third, we estimate parameters using population-level data–a key strategy to address challenges studying early-onset cancers, for which the absolute risks remain low and individual data remain sparse. We are thus unable to fully capture the heterogeneity of carcinogenesis timelines across individuals, and likely underestimate the model uncertainty, as shown by narrow 95% confidence intervals (Tables 1 and 2). These limitations should be considered when interpreting the results.

This work also has several strengths. We provide detailed estimates of carcinogenesis timelines for three key cancers (breast, colorectal, and thyroid), including key differences between early-onset and late-onset cases and changes in early-onset timelines across recent birth cohorts. Consistent with reported early-onset incidence increases, we find accelerated early-onset carcinogenesis timelines in more recent cohorts; we also identify key changes, which can inform early-onset cancer etiology and intervention. Methodologically, we introduce a new approach to estimating the carcinogenesis timeline including intermediate transitions. As it requires population-level data alone, our approach is applicable to early-onset cancers, which are challenging to study due to data sparsity. Moreover, by quantifying both final and intermediate transitions, our modeling framework introduces new avenues for further investigations (e.g., to test how lifestyle and environmental exposures affect different stages of carcinogenesis). Future work could incorporate relevant exposure data to help pinpoint which exposures may have contributed to the accelerated carcinogenesis progression among young adults in recent years and inform early cancer prevention.

## Data availability statement

The data that support the findings of this study are publicly available from the Surveillance, Epidemiology, and End Results Program at https://seer.cancer.gov/data-software/, and from the National Health and Nutrition Examination Survey at https://wwwn.cdc.gov/nchs/nhanes/. Further information is available from the corresponding author upon request.

## Acknowledgement

We would like to thank The Mailman School of Public Health for access to high-performance computing resources.

