## Supplementary Materials for "Estimating the carcinogenesis timelines in early-onset versus late-onset cancers and changes across birth cohorts"

### Model derivation

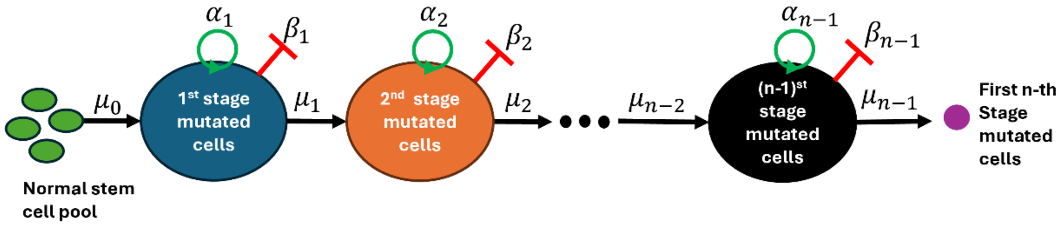


*Figure S1: Schematic of a general n-stage clonal expansion model. The incidence is defined as the appearance of the first n-th stage mutated cell.*

Here, we provide a general derivation for the three classes of model (A, B, and C) presented in the main text. As illustrated in Figure S1, we assume each cell at each stage $i = 1,2,3,\dots,n-1$ can go through one (and only one) of the following events at an infinitesimally small time interval $\Delta t$:

1. Birth: Divide into two identical cells at a rate $\alpha_i$ with the probability $\alpha_i \Delta t + O(\Delta t^2)$,
2. Death: Die at a rate $\beta_i$ with the probability $\beta_i \Delta t + O(\Delta t^2)$,
3. Mutation: Mutate at a rate $\mu_i$ with the probability $\mu_i \Delta t + O(\Delta t^2)$,
4. Stay unchanged with the probability $1 - (\alpha_i + \beta_i + \mu_i)\Delta t + O(\Delta t^2)$.

We assume that the number of stem cells stays constant ($N_0$), therefore we do not consider any birth or death for the normal stem cell pool. Let $I_i(t)$ denote the number of cells in stages $i = 0,1,2,\dots,n$ at time $t$. Here, survival is defined as the probability of not having any n-th stage mutated cells at time $t$ given various initial conditions. Hence, we use the following probability functions

$$P_i(t) = \Pr(I_n(t) < 1 \mid I_0(t) = N_0, I_1(t) = a_1, I_2(t) = a_2, \cdots, I_n(t) = a_n), \qquad \text{(S1)}$$

where, $(a_1, a_2, \cdots, a_n)$ is equal to $(0,0,\cdots,0)$, $(1,0,\cdots,0)$, $(0,1,\cdots,0)$, $\cdots$, $(0,0,\cdots,1,\cdots,0)$, $\cdots$, $(0,0,\cdots,1)$ when $i$ is $0,1,2,\cdots,j,\cdots,n$, respectively. Notice that based on this definition $P_n(t) = 0$, so eq.S1 introduces $n$ nonzero survival probabilities. Note that while survival probabilities can be defined under infinitely many initial conditions, we restrict our definition to this specific set for convenience. This is justified because any survival probability with one or more integers $a_i > 1$ can be expressed as a finite product of survival probabilities corresponding to the cases defined above.

Another necessary assumption is that the cells at each stage are identical and independent, and their transition probabilities are time homogeneous. The process shown in Figure S1 with the assumption of time homogeneity is a Markov process (i.e., the future state depends only on the present state, not on the sequence of events that preceded it), therefore we can utilize Chapman-Kolmogrov's law in the derivation phase. This law states that for a stochastic Markov process with the random variable $I(t)$ and an event A, we can write:

$$\Pr(I(t+s) \in A \mid I(0) = a) = \sum_{k=0}^{\infty} \Pr(I(t+s) \in A \mid I(s) = k) \Pr(I(s) = k \mid I(0) = a). \quad \text{(S2)}$$

For $i = 0$, assuming $\Delta t$ is a sufficiently small time interval (i.e., $O(\Delta t^2) \approx 0$), we have

$$\begin{aligned}
&P_0(t+\Delta t) = \Pr(I_n(t+\Delta t) < 1 \mid I_0(0) = N_0, I_1(0) = 0, \cdots, I_n(0) = 0) \\
&= \sum_{k_0,k_1,k_2,\cdots,k_n} [\Pr(I_n(t+\Delta t) < 1 \mid I_0(\Delta t) = N_0 + k_0, I_1(\Delta t) = k_1, \cdots, I_n(\Delta t) = k_n) \times \\
&\quad Pr(I_0(\Delta t) = N_0 + k_0, I_1(\Delta t) = k_1, \cdots, I_n(\Delta t) = k_n \mid I_0(0) = N_0, I_1(0) = 0, \cdots, I_n(0) = 0)] \\
&= \Pr(I_n(t+\Delta t) < 1 \mid I_0(\Delta t) = N_0, I_1(\Delta t) = 1, \cdots, I_n(\Delta t) = 0)\mu_0 N_0 \Delta t + \\
&\quad \Pr(I_n(t+\Delta t) < 1 \mid I_0(\Delta t) = N_0, I_1(\Delta t) = 0, \cdots, I_n(\Delta t) = 0)(1 - \mu_0 N_0 \Delta t) \\
&= \mu_0 N_0 \Delta t P_0(t) P_1(t) + P_0(t) - \mu_0 N_0 \Delta t P_0(t).
\end{aligned}$$

Here, we used eq.S2 for the first equality. The infinite summation above turns into the sum of two probabilities in the second equality since the normal cells can only mutate to the next stage or stay unchanged, making the rest of the terms in the summation zero. Also, because we have $N_0$ normal stem cells at the beginning, the mutation rate $\mu_0$ is multiplied by $N_0$. The last equality is acquired by utilizing the time homogeneity assumption (i.e., the probabilities in $[0, t]$ and $[\Delta t, t + \Delta t]$ are equivalent) and the independence assumption allowing us to decompose the first probability to $P_0(t)P_1(t)$. To finish this step of derivation, we only need to subtract $P_0(t)$ from both sides and divide by $\Delta t$. Taking the limit as $\Delta t \to 0$ yields:

$$\frac{dP_0(t)}{dt} = -\mu_0 N_0 P_0(t)\big(1 - P_1(t)\big). \quad \text{(S3)}$$

For $i = j$, and sufficiently small $\Delta t$ we can write

$$\begin{aligned}
&P_j(t+\Delta t) = \Pr(I_n(t+\Delta t) < 1 \mid I_0(0) = N_0, \cdots, I_j(0) = 1, \cdots, I_n(0) = 0) \\
&= \sum_{k_0,k_1,\cdots k_j,\cdots,k_n} \big[\Pr(I_n(t+\Delta t) < 1 \mid I_0(\Delta t) = N_0 + k_0, \cdots, I_j(\Delta t) = k_j, \cdots, I_n(\Delta t) = k_n\big) \times \\
&\quad Pr\big(I_0(\Delta t) = N_0 + k_0, \cdots, I_j(\Delta t) = k_j, \cdots, I_n(\Delta t) = k_n \,\big|\, I_0(0) = N_0, \cdots, I_j(0) = 1, I_n(0) = 0)]
\end{aligned}$$

$= \Pr(I_n(t+\Delta t) < 1 \mid I_0(\Delta t) = N_0, \cdots, I_j(\Delta t) = 1, I_{j+1}(\Delta t) = 1, \cdots, I_n(\Delta t) = 0)\mu_j \Delta t +$
$\Pr(I_n(t+\Delta t) < 1 \mid I_0(\Delta t) = N_0, \cdots, I_j(\Delta t) = 2, \cdots, I_n(\Delta t) = 0)\alpha_j \Delta t +$
$\Pr(I_n(t+\Delta t) < 1 \mid I_0(\Delta t) = N_0, \cdots, I_j(\Delta t) = 0, \cdots, I_n(\Delta t) = 0)\beta_j \Delta t +$
$\Pr(I_n(t+\Delta t) < 1 \mid I_0(\Delta t) = N_0, \cdots, I_j(\Delta t) = 1, \cdots, I_n(\Delta t) = 0)(1 - \mu_j \Delta t - \alpha_j \Delta t - \beta_j \Delta t)$
$= \mu_j \Delta t P_j(t) P_{j+1}(t) + \alpha_j \Delta t P_j(t)^2 + \beta_j \Delta t + (1 - \mu_j \Delta t - \alpha_j \Delta t - \beta_j \Delta t) P_j(t).$

This time there are four nonzero terms associated with birth, death, mutation and no change for the j-th stage mutated cells. By subtracting $P_j(t)$ from both sides and dividing by $\Delta t$ and taking the limit as $\Delta t \to 0$, we get the following for $j = 1, \cdots, n-2$.

$$\frac{dP_j}{dt} = \beta_j - (\mu_j + \alpha_j + \beta_j) P_j(t) + \mu_j P_j(t) P_{j+1}(t) + \alpha_j P_j(t)^2. \tag{S4}$$

Given that $P_n(t) = 0$, as mentioned before, for $j = n-1$ according to eq.S4 we have:

$$\frac{dP_{n-1}}{dt} = \beta_{n-1} - (\mu_{n-1} + \alpha_{n-1} + \beta_{n-1}) P_{n-1}(t) + \alpha_{n-1} P_{n-1}(t)^2. \tag{S5}$$

We consider that survival is always guaranteed at $t = 0$, and this provides our system of ODEs with the respective initial conditions $P_i(0) = 1$.
In order to simultaneously solve for the hazard (i.e., $h_0(t) = -\frac{d}{dt} \ln P_0(t)$) we need to incorporate the first derivative of $P_i(t)$ for $i = 1,2, \cdots, n-1$ in the system. This will give us the following ODEs:

$$\frac{dP_0(t)}{dt} = -\mu_0 N_0 P_0(t)(1 - P_1(t)),$$

$$\frac{dh_0(t)}{dt} = -\mu_0 N_0 (P_1(t))',$$

$$\frac{dP_j(t)}{dt} = \beta_j - (\alpha_j + \beta_j + \mu_j) P_j(t) + \alpha_j \left(P_j(t)\right)^2 + \mu_j P_j(t) P_{j+1}(t),$$

$$\frac{d\left(P_j(t)\right)'}{dt} = -(\alpha_j + \beta_j + \mu_j)\left(P_1(t)\right)' + \mu_j \left(P_j(t)\right)' P_{j+1}(t) + \mu_j P_j(t) \left(P_{j+1}(t)\right)' + 2\alpha_j P_j(t) \left(P_j(t)\right)',$$

$$\frac{dP_{n-1}(t)}{dt} = \beta_{n-1} - (\alpha_{n-1} + \beta_{n-1} + \mu_{n-1}) P_{n-1}(t) + \alpha_{n-1} (P_{n-1}(t))^2,$$

$$\frac{d(P_{n-1}(t))'}{dt} = -(\alpha_{n-1} + \beta_{n-1} + \mu_{n-1})\left(P_{n-1}(t)\right)' + 2\alpha_{n-1} P_{n-1}(t)(P_{n-1}(t))',$$

for $j = 1, \cdots, n-2$. As for the initial conditions, we have $P_i(0) = 1$ (for $i = 0,1, \cdots, n-1$), $h_0(0) = 0$, $\left(P_j(t)\right)'|_{t=0} = 0$ (for $j = 1, \cdots, n-2$) and $(P_{n-1}(t))'|_{t=0} = -\mu_{n-1}$. The initial conditions for the hazard and first derivatives are simply acquired by evaluating the right hand side of eqs.S3-S5 at $t = 0$.

Setting $n = 1, 2, 3$ in the above set of ODEs gives model class (A), (B), and (C) presented in the main text, respectively. Note for class (A), $P_1^{(A)}(t) = 0$; hence we have the simplified Eq. 9 in the main text.
For the MSCE-T model, since $P_n(t) \neq 0$, the derivation entails the use of a moment-generating PDE to obtain a closed form replacing $P_n(t)$. For more details, refer to Mohammad Mirzaei et al. 2024 [1].

**Data sources and processing.** Cancer incidence data and tumor size data were sourced from the Surveillance, Epidemiology, and End Results (SEER) program, combining data released in April 2018 (for cases diagnosed during 1973-2015) and in April 2024 (for cases diagnosed during 2000-2021) [2, 3]. Briefly, we used the International Classification of Diseases for Oncology (ICD-O) codes to filter out the three cancer types: 1) C50.0-C50.9 for breast cancer (BrC); 2) C18.0-C18.9, C19.9, and C20.9 for colorectal cancer (CRC); and 3) C73.9 for thyroid cancer (ThC). We divide the data into 5-year birth cohorts and stratify them by sex. Tumor-size-at-diagnosis were reported under 10-digit EOD (1988-2003) or CS tumor size (2004-2021) and describe the largest dimension, or the diameter of the primary tumor, at the time of diagnosis; we estimated tumor size at detection for years without SEER data (1973-1987), using linear extrapolation.

Normal mammary stem cells are in a quiescent state (i.e., inactive) until puberty [4]. Hence, for all analyses related to BrC, we solve the ODEs with a delay corresponding to age at menarche, as done in [1]. Data on age at menarche were obtained from the National Health and Nutrition Examination Survey (NHANES) reproductive health questionnaires [5]. We combined data across survey cycles from 1999–2000 through 2017–2018 and adjusted the sample weights according to NHANES guidelines. Limiting the analysis to respondents born between 1930 and 1994, and to those who reported menarcheal ages between 8 and 20 years, we retained a total of 21,830 individuals. For each 5-year birth cohort (e.g., 1930–1934 through 1990–1994), we calculated the weighted mean age at menarche and its 95% confidence interval using the "survey" package in R [6].

Table S1. Estimated age at menarche by birth cohort

| Cohort | Age at menarche (in years) |
|---|---|
| 1930-1934 | 12.92 (CI: 12.8-13.03) |
| 1935-1939 | 12.97 (CI: 12.87-13.08) |
| 1940-1944 | 12.75 (CI: 12.63-12.88) |
| 1945-1949 | 12.67 (CI: 12.56-12.78) |
| 1950-1954 | 12.69 (CI: 12.57-12.80) |
| 1955-1959 | 12.72 (CI: 12.61-12.83) |
| 1960-1964 | 12.86 (CI: 12.75-12.97) |
| 1965-1969 | 12.76 (CI: 12.66-12.87) |
| 1970-1974 | 12.62 (CI: 12.52-12.72) |
| 1975-1979 | 12.51 (CI: 12.41-12.60) |
| 1980-1984 | 12.52 (CI: 12.43-12.61) |
| 1985-1989 | 12.49 (CI: 12.4-12.57) |
| 1990-1994 | 12.35 (CI: 12.24-12.45) |

**Parameter estimation**. Parameter estimation for each cancer type and birth cohort is carried out 500 times using MATLAB's HGA toolbox, as done in [1]. We filter the resulting parameter sets by excluding the outliers based on their associated data fit costs and use the remaining for our analyses.
